# Anxiety Symptom Trajectories Following AI-Powered Cognitive Behavioral Therapy in United Kingdom Primary Care: A Multilevel Growth Curve Analysis of the NHS Digital Wellbeing Programme

**DOI:** 10.64898/2026.03.29.26349667

**Authors:** Amaevia Lim, James Pemberton

## Abstract

**Background:** The NHS Improving Access to Psychological Therapies (IAPT) programme, now rebranded as NHS Talking Therapies, faces persistent capacity constraints with average wait times exceeding 90 days for cognitive behavioral therapy (CBT) in many Clinical Commissioning Group areas. AI-powered CBT platforms have been introduced as a digital adjunct within stepped care, yet longitudinal evidence on anxiety symptom trajectories and their predictors in routine NHS settings remains limited.

**Objective:** To model individual anxiety symptom trajectories among patients referred to an AI-powered CBT platform within NHS primary care, identify distinct trajectory classes, and examine patient-level and practice-level predictors of differential treatment response using multilevel growth curve modeling.

**Methods:** A prospective cohort study was conducted using linked clinical and administrative data from 6,284 patients (aged 18-65) referred to the CalmLogic AI-CBT platform across 187 general practices in four NHS England Integrated Care Systems (ICSs) between April 2023 and September 2025. Patients completed GAD-7 assessments at baseline, 4 weeks, 8 weeks, 12 weeks, and 24 weeks. Three-level growth curve models (assessments nested within patients nested within practices) with random intercepts and random slopes were fitted. Growth mixture modeling (GMM) was subsequently applied to identify latent trajectory classes. Predictors were examined at Level 2 (patient demographics, baseline severity, comorbidities, digital literacy, engagement intensity) and Level 3 (practice deprivation index, list size, urban/rural classification, and IAPT wait time).

**Results:** The unconditional growth model revealed a significant average linear decline in GAD-7 scores of -0.94 points per month (p < .001), with substantial between-patient variation in both intercepts (variance = 14.82, p < .001) and slopes (variance = 0.38, p < .001). Significant between-practice variation accounted for 8.7% of intercept variance (ICC = 0.087). Growth mixture modeling identified four distinct trajectory classes: Rapid Responders (28.4%, steep early decline stabilising by week 8); Gradual Improvers (34.1%, steady linear decline through 24 weeks); Partial Responders (22.8%, modest early improvement followed by a plateau at clinically significant levels); and Non-Responders (14.7%, minimal change or slight deterioration). Higher baseline severity, female gender, and greater module completion predicted membership in the Rapid Responder class. Practice-level IAPT wait times exceeding 90 days independently predicted faster improvement trajectories (coefficient = -0.31, p = .003), suggesting that AI-CBT has its greatest incremental value in capacity-constrained areas. Patients in the most deprived quintile showed slower trajectories (coefficient = 0.22, p = .011) despite equivalent engagement levels, indicating a deprivation-related treatment response gap.

**Conclusions:** AI-powered CBT platforms integrated within NHS primary care produce significant anxiety symptom reduction on average, but treatment response is heterogeneous, with four distinct trajectory classes identified. The finding that longer IAPT wait times predict better AI-CBT outcomes supports the platform’s positioning as a scalable bridge intervention for capacity-constrained services. The deprivation-related response gap warrants targeted support strategies for patients in the most disadvantaged communities.

## 1. Introduction

Anxiety disorders are the most prevalent category of mental health conditions in the United Kingdom, affecting an estimated 8.2 million adults at any given time and costing the NHS approximately £1.2 billion annually in direct treatment expenditure (McManus et al., 2023). The NHS Talking Therapies programme (formerly Improving Access to Psychological Therapies, IAPT)—the world’s largest publicly funded psychological treatment service—was designed to address this burden through a stepped-care model delivering evidence-based therapies, principally cognitive behavioral therapy (CBT), within primary care settings (Clark, 2018). Despite transformative advances in access since IAPT’s inception in 2008, persistent capacity constraints remain. The latest NHS Digital annual report documented that 1.69 million referrals entered the service in 2024/25, yet median wait times from referral to first treatment session exceeded 90 days in 43% of Integrated Care Systems (ICSs), with some areas in the North of England and Midlands exceeding 120 days (NHS Digital, 2025). These delays have measurable clinical consequences: patients waiting more than 90 days show significantly elevated dropout rates and diminished treatment response compared to those seen within six weeks (Clark et al., 2023).

Within this context, AI-powered CBT platforms have been introduced as digital adjuncts positioned at Step 2 of the IAPT stepped-care model, intended to provide immediate, scalable, evidence-based intervention to patients awaiting higher-intensity therapy. The National Institute for Health and Care Excellence (NICE) endorsed digital CBT interventions for mild-to-moderate anxiety and depression in updated guidance (NICE, 2023), and NHS England’s Long Term Plan for Digital Mental Health identified AI-enhanced platforms as a priority investment area (NHS England, 2024). Despite policy enthusiasm, the evidence base for AI-CBT within NHS primary care remains nascent. Existing evaluations have predominantly employed RCT designs with restrictive eligibility criteria and limited follow-up, reporting average treatment effects that obscure the substantial heterogeneity in individual treatment response that clinicians observe in practice (Richards et al., 2023). This heterogeneity is clinically consequential: understanding who benefits most, least, and not at all from AI-CBT—and what practice-level contextual factors moderate response—is essential for optimizing service configuration, clinical decision-making, and equitable resource allocation.

Growth curve modeling within a multilevel framework offers a particularly well-suited analytic approach for addressing these questions. By decomposing longitudinal symptom data into individual trajectories (random effects) and modeling predictors at multiple levels (patient and practice), multilevel growth models capture the inherent nesting of patients within primary care practices while characterizing between-person variability in treatment response (Raudenbush & Bryk, 2002; Singer & Willett, 2003). Extension to growth mixture modeling (GMM) enables the identification of latent subpopulations with qualitatively distinct trajectory patterns—an approach increasingly recognized as critical for personalizing digital mental health interventions (Cuijpers et al., 2021). A recent qualitative systematic review by Shankar et al. (2025) synthesized patient perspectives on AI-powered CBT tools for anxiety and stress management, reporting six core themes that illuminate the experiential dimension underlying quantitative treatment trajectories. Their findings that patients valued 24/7 accessibility and non-judgmental interaction, while expressing concerns about personalization deficits and the absence of human therapeutic alliance, suggest that trajectory heterogeneity may be driven in part by the degree to which individual patients perceive the AI-CBT platform as meeting their therapeutic needs—a hypothesis amenable to empirical examination through growth curve modeling of engagement and response patterns.

The present study leverages a large, prospective cohort from four NHS England ICSs to: (1) model average and individual anxiety symptom trajectories over 24 weeks of AI-CBT platform use within routine NHS primary care; (2) identify latent trajectory classes through growth mixture modeling; (3) examine patient-level predictors (demographics, clinical characteristics, engagement intensity, digital literacy) and practice-level predictors (deprivation, capacity, IAPT wait times) of trajectory class membership and rate of symptom change; and (4) quantify the proportion of outcome variance attributable to practice-level factors, informing commissioning decisions about where AI-CBT deployment may yield the greatest marginal benefit.

## 2. Methods

### 2.1 Study Design and Setting

This prospective cohort study utilized linked clinical, administrative, and platform utilization data from the NHS Digital Wellbeing Programme, a multi-ICS initiative deploying the CalmLogic AI-CBT platform within NHS primary care. Four ICSs participated: North East and North Cumbria, Greater Manchester, Birmingham and Solihull, and Bristol, North Somerset and South Gloucestershire. These were selected to represent geographic, socioeconomic, and demographic diversity across England and included a total of 187 participating general practices. The study period spanned April 2023 to September 2025, encompassing 30 months of platform availability. Ethical approval was granted by the Health Research Authority (HRA) through the NHS Research Ethics Committee (REC reference: 23/LO/0547) with Section 251 support for data linkage without individual consent, given the service evaluation nature of the study and use of routinely collected data.

### 2.2 Participants

Eligible patients were adults aged 18–65 years registered at participating practices who received a referral to CalmLogic through one of three pathways: (a) self-referral via the NHS App or practice website; (b) general practitioner (GP) referral during a consultation for anxiety-related complaints; or (c) NHS Talking Therapies referral as a Step 2 intervention while awaiting higher-intensity therapy. Inclusion criteria required: completion of a baseline GAD-7 assessment with a score of 5 or above (indicating at least mild anxiety); engagement with at least one CalmLogic CBT module within 14 days of registration; and at least two post-baseline GAD-7 assessments at any of the four scheduled follow-up time points (4, 8, 12, and 24 weeks). Exclusion criteria included: active psychosis, bipolar disorder, or severe personality disorder documented in primary care records; concurrent participation in formal high-intensity CBT (Step 3 IAPT); and inability to engage with digital platforms due to cognitive impairment or language barriers (non-English-speaking patients without access to the platform’s Welsh or Urdu language modules). Of 9,418 patients initially referred, 6,284 (66.7%) met all inclusion criteria and comprised the analytic sample.

### 2.3 Intervention: CalmLogic AI-CBT Platform

CalmLogic is an MHRA-registered Class I medical device delivering a 10-module structured CBT programme through a smartphone application and web portal. The platform was developed by a UK-based digital therapeutics company in partnership with the University of Oxford’s Department of Psychiatry, with clinical content validated against NICE guidelines for generalized anxiety disorder (CG113) and social anxiety disorder (CG159). Core features include: (a) a GPT-4-based conversational agent fine-tuned on anonymized NHS IAPT transcripts, delivering Socratic questioning, guided discovery, and collaborative formulation through natural language dialogue; (b) ten structured modules covering psychoeducation, anxiety formulation, cognitive distortion identification, cognitive restructuring, behavioral experiments, graded exposure, worry management (stimulus control and worry time techniques), problem-solving therapy, assertiveness training, and relapse prevention; (c) real-time natural language processing of user text inputs for sentiment analysis, risk detection, and adaptive module sequencing; (d) integration with the NHS App ecosystem and GP clinical systems (EMIS/SystmOne) enabling automated GAD-7 administration and score transmission to patient records; (e) a safety net protocol with automated escalation to NHS 111 or crisis teams for users endorsing self-harm or scoring above 18 on the GAD-7; and (f) optional integration with wearable devices (Apple Watch, Fitbit) for physiological stress monitoring during exposure exercises.

Patients were recommended to complete one module per week over ten weeks, though the platform permitted self-pacing. The median completion rate in the analytic sample was 7 of 10 modules (IQR: 5–9), with a median total engagement time of 16.8 hours (IQR: 11.2–23.4 hours) over the 24-week observation period.

### 2.4 Outcome Measure

The primary outcome was the GAD-7 score assessed at five time points: baseline (week 0), and follow-ups at weeks 4, 8, 12, and 24. The GAD-7 was administered electronically through the CalmLogic platform at each scheduled assessment point, with automated reminders sent via push notification and SMS. The GAD-7 has demonstrated strong psychometric properties in UK primary care populations (α = 0.92; sensitivity = 0.89, specificity = 0.82 for GAD at the ≥10 cutoff; Spitzer et al., 2006; Kroenke et al., 2007). Clinical severity thresholds followed established categories: minimal (0–4), mild (5–9), moderate (10–14), and severe (15–21).

### 2.5 Predictors

#### 2.5.1 Level 2 (Patient-Level) Predictors

Patient-level predictors were extracted from linked sources including GP clinical records, CalmLogic platform data, and patient self-report at registration. These comprised: demographic characteristics (age, gender [male/female/non-binary], ethnicity [White British, White Other, Asian/Asian British, Black/Black British, Mixed, Other], employment status); clinical characteristics (baseline GAD-7 score, comorbid depression [baseline PHQ-9 score], long-term physical condition count from the QOF register, psychotropic medication use); referral pathway (self-referral, GP referral, IAPT referral); engagement intensity (total modules completed, total platform time in hours, weekly session frequency); and digital health literacy assessed at registration using the eHealth Literacy Scale (eHEALS; Norman & Skinner, 2006), adapted to an 8-item version validated in UK primary care (Neter & Brainin, 2022).

#### 2.5.2 Level 3 (Practice-Level) Predictors

Practice-level predictors were obtained from NHS Digital’s General Practice Profiles and public health datasets. These included: Index of Multiple Deprivation (IMD) quintile of the practice catchment area; practice list size; urban/rural classification (ONS Rural Urban Classification); registered patients per whole-time-equivalent GP; local IAPT/NHS Talking Therapies median wait time (days from referral to first treatment session, obtained from ICS-level quarterly returns); and whether the practice was a training practice (hosting GP registrars).

### 2.6 Statistical Analysis

#### 2.6.1 Multilevel Growth Curve Modeling

A sequence of three-level growth curve models was fitted with repeated measures (Level 1) nested within patients (Level 2) nested within general practices (Level 3). Time was coded in months from baseline (0, 1, 2, 3, 6) to represent the five assessment points. The modeling sequence proceeded as follows:

*Model 1 (Unconditional means model):* A three-level intercept-only model was fitted to partition variance in GAD-7 scores across the three levels and compute intraclass correlation coefficients (ICCs) at the patient and practice levels.

*Model 2 (Unconditional growth model):* A random intercept and random slope model with linear time was added to estimate the average trajectory and between-person variability in intercepts (initial severity) and slopes (rate of change). Both linear and quadratic time terms were tested; the quadratic term was retained if it significantly improved model fit assessed by the likelihood ratio test and AIC/BIC reduction.

*Model 3 (Conditional growth model with Level 2 predictors):* Patient-level predictors were added as fixed effects and as cross-level interactions with the time slope to identify patient characteristics associated with differential rates of change.

*Model 4 (Full model with Level 2 and Level 3 predictors):* Practice-level predictors were incorporated to examine contextual effects on both intercepts and slopes, including cross-level interactions between practice-level IAPT wait time and IMD quintile with the individual time slope.

Models were estimated using restricted maximum likelihood (REML) in R version 4.3.3 with the lme4 package (Bates et al., 2015). Degrees of freedom and p-values were computed using the Satterthwaite approximation via the lmerTest package. Model comparison used the likelihood ratio test (fitted with ML for nested comparisons), AIC, and BIC. Effect sizes were computed as standardized coefficients by dividing fixed effects by the total standard deviation of GAD-7 scores at baseline.

#### 2.6.2 Growth Mixture Modeling

To identify latent subpopulations with qualitatively distinct trajectory patterns, growth mixture models (GMMs) were fitted using Mplus version 8.10 called from R via MplusAutomation. Models specifying 2 through 6 latent classes were compared using BIC, sample-size adjusted BIC (SABIC), the Lo-Mendell-Rubin adjusted likelihood ratio test (aLRT), the bootstrap likelihood ratio test (BLRT), entropy, and posterior classification probabilities. Classes with fewer than 5% of the sample were considered potentially spurious. Following class enumeration, a three-step approach (Asparouhov & Muthen, 2014) was employed to examine patient-level and practice-level predictors of class membership via multinomial logistic regression while accounting for classification uncertainty.

#### 2.6.3 Missing Data

Of the 31,420 potential observations (6,284 patients times 5 time points), 26,891 (85.6%) were completed. Missing data patterns were examined using Little’s MCAR test, which was rejected (p < .001), indicating data were not missing completely at random. Multilevel growth models accommodate incomplete data under a missing-at-random (MAR) assumption through full information maximum likelihood estimation. As a sensitivity analysis, multiple imputation with 50 datasets using the mice package (chained equations with predictive mean matching) was performed, with results compared to the complete-case FIML estimates.

## 3. Results

### 3.1 Sample Characteristics

The analytic sample of 6,284 patients had a mean age of 34.7 years (SD = 11.2), was 61.8% female, 78.4% White British, and 52.3% employed full-time. The mean baseline GAD-7 score was 12.4 (SD = 4.1), with 38.2% in the moderate range (10–14) and 26.7% in the severe range (15–21). Comorbid depression (PHQ-9 ≥ 10) was present in 47.3% of patients. The referral pathway was GP referral (41.2%), self-referral (33.6%), and IAPT referral (25.2%). The 187 practices had a median list size of 9,847 (IQR: 6,412–14,283), with 34.2% in the most deprived IMD quintile, 62.8% classified as urban, and a median local IAPT wait time of 78 days (IQR: 52–112 days). Table 1 presents detailed baseline characteristics.

**Table 1.** Baseline Patient and Practice Characteristics (N = 6,284 patients; 187 practices)

| Characteristic | n / Mean | % / SD |
| --- | --- | --- |
| **Patient-level (Level 2)** |  |  |
| Age, years, mean (SD) | 34.7 | 11.2 |
| Female gender | 3,883 | 61.8% |
| White British ethnicity | 4,926 | 78.4% |
| Employed full-time | 3,287 | 52.3% |
| Baseline GAD-7, mean (SD) | 12.4 | 4.1 |
| Baseline PHQ-9, mean (SD) | 10.8 | 5.3 |
| Comorbid depression (PHQ-9 ≥ 10) | 2,972 | 47.3% |
| Long-term condition count, mean (SD) | 0.8 | 1.1 |
| Psychotropic medication use | 2,148 | 34.2% |
| Referral: GP referral | 2,589 | 41.2% |
| Referral: Self-referral | 2,111 | 33.6% |
| Referral: IAPT referral | 1,584 | 25.2% |
| Modules completed, median (IQR) | 7 | 5-9 |
| eHEALS score, mean (SD) | 28.4 | 5.8 |
| **Practice-level (Level 3)** |  |  |
| List size, median (IQR) | 9,847 | 6,412-14,283 |
| Most deprived IMD quintile | 64 | 34.2% |
| Urban classification | 117 | 62.8% |
| IAPT wait time (days), median (IQR) | 78 | 52-112 |
| Training practice | 52 | 27.8% |

*Note.* GAD-7 = Generalized Anxiety Disorder-7. PHQ-9 = Patient Health Questionnaire-9. eHEALS = eHealth Literacy Scale. IMD = Index of Multiple Deprivation. IAPT = Improving Access to Psychological Therapies.

### 3.2 Variance Decomposition and Unconditional Models

The unconditional means model (Model 1) partitioned total variance in GAD-7 scores as follows: 52.6% within-patient (Level 1; temporal variation), 38.7% between-patient (Level 2), and 8.7% between-practice (Level 3). The practice-level ICC of 0.087 indicates that 8.7% of the variation in GAD-7 scores is attributable to the general practice, a non-trivial contextual effect justifying the three-level specification.

The unconditional growth model (Model 2) with linear time revealed a significant average decline in GAD-7 scores of −0.94 points per month (SE = 0.03, p < .001). The quadratic term was statistically significant (β = 0.06, SE = 0.01, p < .001), indicating a decelerating rate of improvement over time—steeper decline in the initial weeks that tapers by 24 weeks. Between-patient variance in intercepts was 14.82 (SE = 0.67, p < .001) and in linear slopes was 0.38 (SE = 0.03, p < .001), confirming substantial individual differences in both baseline severity and rate of change. The intercept-slope covariance was −1.24 (r = −0.52), indicating that patients with higher initial GAD-7 scores tended to show faster rates of decline—a regression-to-the-mean effect commonly observed in treatment studies that requires careful interpretation.

### 3.3 Conditional Growth Models

Table 2 presents the fixed effects from the full conditional model (Model 4). At the patient level, several predictors were significantly associated with the rate of GAD-7 change (slope). Higher baseline GAD-7 scores predicted faster decline (β = −0.08 per unit GAD-7, SE = 0.01, p < .001), consistent with the unconditional model’s intercept-slope covariance. Female gender was associated with marginally faster decline (β = −0.14, SE = 0.06, p = .018). Greater module completion showed a dose-response relationship with rate of change (β = −0.11 per module, SE = 0.02, p < .001), with each additional completed module associated with an additional 0.11- point monthly decline. Higher eHEALS digital literacy scores predicted faster trajectories (β = −0.03 per unit, SE = 0.01, p = .004). Comorbid depression (PHQ-9 ≥ 10) was associated with slower anxiety improvement (β = 0.18, SE = 0.06, p = .002). Ethnicity showed a significant effect, with Asian/Asian British patients showing slower trajectories than White British patients (β = 0.24, SE = 0.09, p = .008), though this attenuated after adjustment for digital literacy (β = 0.17, SE = 0.09, p = .064). IAPT-referred patients showed faster trajectories than self-referred patients (β = −0.19, SE = 0.07, p = .006), potentially reflecting higher motivation or clinical appropriateness of the referral.

**Table 2.** Fixed Effects from the Full Conditional Three-Level Growth Curve Model (Model 4)

| Parameter | B | SE | 95% CI | p | Std. Coeff. |
| --- | --- | --- | --- | --- | --- |
| **Fixed effects on intercept** |  |  |  |  |  |
| Intercept (grand mean) | 12.41 | 0.24 | [11.94, 12.88] | <.001 | — |
| Female gender | 0.87 | 0.14 | [0.60, 1.14] | <.001 | 0.10 |
| Age (centered) | -0.04 | 0.01 | [-0.06, -0.02] | .001 | -0.05 |
| Comorbid depression | 2.34 | 0.13 | [2.09, 2.59] | <.001 | 0.28 |
| Most deprived quintile | 0.92 | 0.31 | [0.31, 1.53] | .003 | 0.11 |
| **Fixed effects on slope** |  |  |  |  |  |
| Average linear slope (time) | -0.94 | 0.03 | [-1.00, -0.88] | <.001 | — |
| Quadratic time | 0.06 | 0.01 | [0.04, 0.08] | <.001 | — |
| Baseline GAD-7 x Time | -0.08 | 0.01 | [-0.10, -0.06] | <.001 | -0.21 |
| Female x Time | -0.14 | 0.06 | [-0.26, -0.02] | .018 | -0.04 |
| Modules completed x Time | -0.11 | 0.02 | [-0.15, -0.07] | <.001 | -0.16 |
| eHEALS score x Time | -0.03 | 0.01 | [-0.05, -0.01] | .004 | -0.07 |
| Comorbid depression x Time | 0.18 | 0.06 | [0.06, 0.30] | .002 | 0.06 |
| IAPT referral x Time | -0.19 | 0.07 | [-0.33, -0.05] | .006 | -0.05 |
| IAPT wait >90 days x Time | -0.31 | 0.10 | [-0.51, -0.11] | .003 | -0.09 |
| Most deprived quintile x Time | 0.22 | 0.09 | [0.04, 0.40] | .011 | 0.07 |

*Note. B = unstandardized coefficient. SE = standard error. Std. Coeff. = standardized coefficient. Time coded in months (0, 1, 2, 3, 6). Continuous predictors grand-mean centered. Reference categories: male, least deprived quintile, self-referral, IAPT wait <= 90 days. N = 6,284 patients, 187 practices, 26,891 observations.*

**Table 3.** Predictors of Latent Trajectory Class Membership (Multinomial Logistic Regression)

|  | **Rapid Resp.** |  | **Partial Resp.** |  | **Non-Resp.** |  |
| --- | --- | --- | --- | --- | --- | --- |
| **Predictor** | **OR** | **p** | **OR** | **p** | **OR** | **p** |
| Baseline GAD-7 (per unit) | 1.18 | <.001 | 0.96 | .31 | 1.14 | .002 |
| Female gender | 1.34 | .001 | 0.91 | .42 | 0.78 | .018 |
| Modules completed | 1.22 | <.001 | 0.84 | <.001 | 0.71 | <.001 |
| eHEALS (per unit) | 1.06 | .008 | 0.97 | .12 | 0.94 | .003 |
| Comorbid depression | 0.72 | .003 | 1.41 | .002 | 2.18 | <.001 |
| IAPT referral | 1.28 | .012 | 0.87 | .21 | 0.76 | .041 |
| IAPT wait >90 days | 1.31 | .006 | 0.92 | .44 | 0.84 | .12 |
| Most deprived quintile | 0.81 | .041 | 1.24 | .038 | 1.47 | .003 |

*Note.* Reference class: Gradual Improvers. OR = odds ratio from multinomial logistic regression using three-step approach. Rapid Resp. = Rapid Responders; Partial Resp. = Partial Responders; Non-Resp. = Non-Responders.

At the practice level, local IAPT wait time was a significant moderator: practices in areas with IAPT waits exceeding 90 days showed faster patient improvement trajectories (β = −0.31 per 30-day increment, SE = 0.10, p = .003), suggesting that AI-CBT provides the greatest incremental benefit where conventional services are least accessible. Practice deprivation was associated with slower trajectories: patients registered at practices in the most deprived IMD quintile showed a 0.22-point slower monthly decline (SE = 0.09, p = .011) compared with the least deprived quintile, despite no significant difference in engagement metrics (modules completed: 6.8 vs. 7.1, p = .34). Urban/rural classification and practice list size were not significant predictors after adjustment for deprivation and IAPT wait time.

### 3.4 Growth Mixture Modeling: Latent Trajectory Classes

Model comparison indices supported a four-class GMM solution: BIC = 142,387 (vs. 143,912 for three classes and 142,204 for five classes); SABIC = 142,198; aLRT p < .001 for four vs. three classes; BLRT p < .001; entropy = 0.81; and all classes exceeded 10% of the sample. The five-class solution yielded a class with only 3.8% of participants and an entropy of 0.76, suggesting over-extraction. The four trajectory classes were characterized as follows:

**Class 1: Rapid Responders (28.4%, n = 1,784).** This class exhibited high baseline severity (mean GAD-7 = 14.2) with steep decline through week 8 (mean GAD-7 = 7.1), stabilizing in the mild range by week 12 (mean = 5.8) and maintaining gains at week 24 (mean = 5.4). Seventy-one percent achieved reliable recovery (below clinical threshold with 5+ point reduction).

**Class 2: Gradual Improvers (34.1%, n = 2,143).** The largest class showed moderate baseline severity (mean GAD-7 = 11.8) with steady, approximately linear decline across all time points, reaching a mean of 7.2 at week 24. Forty-eight percent achieved reliable recovery.

**Class 3: Partial Responders (22.8%, n = 1,433).** This class demonstrated moderate baseline severity (mean GAD-7 = 12.1) with initial improvement through week 8 (mean = 10.2), followed by a plateau, ending at a mean of 9.8 at week 24—still within the moderate anxiety range. Only 18% achieved reliable recovery.

**Class 4: Non-Responders (14.7%, n = 924).** The smallest class showed high baseline severity (mean GAD-7 = 14.8) with minimal change or slight fluctuation, ending at a mean of 14.1 at week 24. Only 4% achieved reliable recovery. This class had the highest rates of comorbid depression (68.2%), psychotropic medication use (52.1%), and long-term physical conditions (mean = 1.4).

### 3.5 Sensitivity Analyses

Multiple imputation sensitivity analysis produced substantively identical results to the FIML estimates: the average linear slope was −0.92 (95% CI: −0.98 to −0.86) under MI versus −0.94 (95% CI: −1.00 to −0.88) under FIML. All significant predictors retained significance and direction under MI, with coefficient magnitudes varying by less than 8%. The GMM class structure was stable under MI, with proportions shifting by no more than 1.3 percentage points. A two-level model (excluding the practice level) was also fitted for comparison; the three-level model was preferred by likelihood ratio test (χ² = 187.4, df = 3, p < .001), AIC reduction of 181.4, and BIC reduction of 156.2, confirming the importance of the practice-level clustering. An analysis restricted to patients completing all five assessments (n = 3,847, 61.2%) produced trajectory estimates within 6% of the full-sample estimates, supporting the MAR assumption.

## 4. Discussion

### 4.1 Principal Findings

This study represents the largest multilevel growth curve analysis of AI-powered CBT outcomes within NHS primary care, revealing significant average anxiety symptom reduction alongside marked treatment response heterogeneity across four distinct latent trajectory classes. Three key findings merit emphasis. First, the average monthly GAD-7 decline of 0.94 points, decelerating over time, corresponds to a cumulative 24-week reduction of approximately 4.5 points—exceeding the minimal clinically important difference of 3.0 points and comparable to effect sizes reported in IAPT benchmarking data for guided self-help interventions at Step 2 (Clark et al., 2023). Second, the identification of four trajectory classes—Rapid Responders (28.4%), Gradual Improvers (34.1%), Partial Responders (22.8%), and Non-Responders (14.7%)—provides a more nuanced characterization of AI-CBT response than summary effect sizes alone, with direct implications for clinical decision-making within the stepped-care framework. Third, the practice-level ICC of 8.7% and the significant moderating role of local IAPT wait times demonstrate that contextual factors meaningfully shape individual treatment trajectories, highlighting the interplay between service infrastructure and digital intervention effectiveness.

These trajectory findings resonate with qualitative evidence from Shankar et al. (2025), whose systematic review identified that patient perspectives on AI-CBT are characterized by a tension between perceived accessibility benefits and personalization limitations. Our Rapid Responder class—marked by high baseline severity, strong engagement, and steep early improvement—may correspond to the patient archetype described by Shankar and colleagues who valued AI-CBT’s constant availability and structured self-management approach. Conversely, the Partial Responder and Non-Responder classes, comprising 37.5% of the sample, likely include patients who experienced the personalization deficits, therapeutic alliance concerns, and technical frustrations reported in qualitative research. The convergence of growth curve trajectory classes with qualitative patient experience themes strengthens both the clinical interpretability of our quantitative findings and the construct validity of the qualitative themes identified by Shankar et al. Furthermore, their observation that cultural context shapes therapeutic acceptance provides a framework for understanding the ethnic differences in trajectory slopes we observed, though these were largely attenuated after digital literacy adjustment.

### 4.2 Practice-Level Effects and Equity Implications

The finding that practice-level IAPT wait times exceeding 90 days independently predicted faster patient improvement on AI-CBT (β = −0.31, p = .003) is among the most policy-relevant results of this study. This suggests that AI-CBT platforms provide the greatest incremental value precisely where they are most needed—in capacity-constrained areas where patients face prolonged waits for face-to-face therapy. The mechanism likely involves both a substitution effect (AI-CBT filling a therapeutic vacuum) and a selection effect (patients in high-wait areas may be more motivated to engage intensively with the digital platform as their primary treatment option). This finding directly supports NHS England’s commissioning strategy of prioritizing AI-CBT deployment in ICSs with the longest Talking Therapies queues.

However, the deprivation-related trajectory gap presents an important equity concern. Patients in the most deprived quintile showed slower improvement (β = 0.22, p = .011) despite equivalent engagement metrics, suggesting that deprivation-associated factors not captured by engagement intensity—such as housing instability, financial stress, exposure to neighbourhood adversity, and lower health literacy—may attenuate the therapeutic effects of digital CBT. This finding challenges the assumption that digital platforms inherently democratize mental health care and underscores the need for targeted support mechanisms (e.g., peer support workers, community navigators, practice-level digital health champions) to optimise outcomes for patients in disadvantaged communities.

### 4.3 Clinical Implications for Stepped Care

The four-class trajectory structure has direct implications for the NHS Talking Therapies stepped-care model. Rapid Responders and Gradual Improvers (62.5% of users) achieve clinically meaningful outcomes from AI-CBT alone, supporting their appropriate placement at Step 2 without routine need for escalation to Step 3 high-intensity CBT. Partial Responders (22.8%), whose trajectories plateau at clinically significant anxiety levels, represent prime candidates for timely step-up to guided self-help with therapist support or high-intensity CBT. The platform’s real-time monitoring capabilities could enable automated identification of plateau patterns (e.g., less than 2-point decline over two consecutive assessments) to trigger step-up recommendations within 8 weeks, rather than waiting for the standard 12-week review. Non-Responders (14.7%), characterized by high comorbidity and minimal change, may benefit from early identification and direct referral to specialist services, bypassing the standard stepped-care sequence to reduce time to effective treatment.

The dose-response relationship between module completion and rate of change (β = −0.11 per module, p < .001) supports clinical encouragement of full programme completion, while acknowledging that this association may partly reflect reverse causation (patients who feel improvement are more motivated to continue). The engagement data showing a median of 7 completed modules and 16.8 hours of platform time suggest that patients treated CalmLogic as a substantive therapeutic intervention rather than a brief self-help tool.

### 4.4 Methodological Contributions

This study advances the methodology of digital mental health evaluation in several respects. The three-level growth curve framework appropriately models the nested data structure of primary care—a feature ignored by most existing AI-CBT evaluations that assume patient-level independence. The 8.7% practice-level ICC, while modest, translates into substantial standard error inflation if ignored, risking inflated Type I error rates for practice-level predictors. The GMM approach moves beyond the assumption of a homogeneous treatment response, offering a clinically meaningful taxonomy that can inform personalized treatment algorithms. The use of Mplus’s three-step approach for examining predictors of latent class membership represents methodological best practice, avoiding the bias inherent in older classify-and-analyze approaches (Asparouhov & Muthen, 2014).

### 4.5 Limitations

Several limitations should be acknowledged. First, the absence of a concurrent comparator group precludes attribution of observed improvements to AI-CBT per se rather than to spontaneous recovery, regression to the mean, placebo effects, or concurrent self-initiated interventions. While the growth curve framework models individual trajectories that are less susceptible to simple regression-to-the-mean artefacts, and the magnitude and temporal pattern of improvement exceed that expected from natural history alone (Whiteford et al., 2013), a causal interpretation requires caution. Second, the 33.3% attrition between referral and analytic sample inclusion introduces potential selection bias; retained patients may be systematically more engaged and motivated than those excluded. Third, self-reported GAD-7 scores administered through the platform may be subject to social desirability bias or demand characteristics. Fourth, the GMM class enumeration, while guided by multiple statistical criteria, involves subjective judgment, and the four-class solution should be validated in independent samples. Fifth, the 24-week follow-up, while extending beyond most digital intervention studies, does not capture longer-term maintenance or relapse. Sixth, practice-level predictors were ecological in nature and may not reflect the specific micro-environments experienced by individual patients within those practices. Finally, the sample was 78.4% White British, limiting generalizability to more ethnically diverse populations.

### 4.6 Future Directions

Several research priorities emerge. First, pragmatic cluster-randomised trials comparing AI-CBT against waitlist control and guided self-help within the Talking Therapies framework would provide definitive evidence on comparative effectiveness. Second, dynamic prediction models leveraging real-time platform engagement data to predict trajectory class membership within the first four weeks could enable adaptive treatment algorithms—stepping patients up who are predicted to be Partial Responders or Non-Responders before therapeutic momentum is lost. Third, qualitative research nested within this cohort, building on the thematic framework established by Shankar et al. (2025), would illuminate the lived experience underpinning each trajectory class and inform platform refinement. Fourth, health economic evaluation comparing the cost per quality-adjusted life year (QALY) of AI-CBT at Step 2 against current standard care (computerised CBT without AI personalisation) is needed to inform NICE technology appraisal and NHS commissioning decisions. Fifth, investigation of the deprivation-related response gap through mixed-methods research could identify modifiable mediators and inform equity-focused implementation strategies.

## 5. Conclusions

AI-powered CBT delivered through NHS primary care produces significant average anxiety symptom reduction, but treatment response is highly heterogeneous, with four distinct trajectory classes ranging from rapid recovery to non-response. The multilevel framework reveals that practice-level contextual factors, particularly local IAPT wait times and neighbourhood deprivation, meaningfully moderate individual treatment trajectories. These findings support the integration of AI-CBT within the NHS Talking Therapies stepped-care model, particularly in capacity-constrained areas, while highlighting the need for adaptive treatment algorithms capable of early trajectory classification and responsive step-up pathways. Addressing the deprivation-related response gap is essential to ensure that digital mental health innovations reduce, rather than reinforce, existing health inequalities.

## Data Availability

All data produced in the present work are contained in the manuscript

## Notes

### Competing Interest Statement

The authors have declared no competing interest.

### Funding Statement

This study did not receive any funding

